# Medical Clinical Minds Meet Artificial Intelligence: Italian Physicians’ Knowledge, Attitudes, and Concordance between Italian Physicians and AI-Generated Diagnoses. A National Cross-Sectional Study

**DOI:** 10.1101/2025.07.30.25332423

**Authors:** Vincenza Cofini, Mario Muselli, Laura Piccardi, Eugenio Benvenuti, Ginevra Di Pangrazio, Martina Mancinelli, Eleonora Cimino, Patrizia Palermo, Emiliano Petrucci, Giovanna Picchi, Loreta Tobia, Arcangelo Barbonetti, Giovambattista Desideri, Leila Fabiani, Maurizio Guido, Franco Marinangeli, Stefano Necozione

## Abstract

**Background:** Artificial Intelligence has increasingly been integrated into clinical practice, yet its adoption and perception among medical professionals remain poorly understood, particularly in the Italian healthcare system. To investigate Italian physicians’ knowledge, attitudes, and clinical concordance with AI-generated diagnostic recommendations, using a validated questionnaire and a clinical scenario processed by ChatGPT.

**Methods:** A national, cross-sectional web-based survey was conducted among 587 Italian physicians using an online validated questionnaire. The first part of the questionnaire assessed self-reported knowledge, prior experience, attitudes, and willingness to adopt AI in medicine. The second part assessed clinical concordance between AI proposals and physicians about clinical cases evaluated by ChatGPT. Results

Most participants reported basic AI knowledge (n=380, 64.8%) and minimal exposure to AI training (18.4%). Only 21.6% reported they used AI in clinical practice, and the most familiar application was diagnostic imaging (35.4% of AI users; 7.7% of the total sample). Major perceived barriers included lack of training (76.7%) and resistance to change (50.9%). In the universal clinical scenario, physicians showed the highest agreement with ChatGPT’s correct diagnosis (mean=4.07) compared to incorrect alternatives (mean=2.57 and 1.82, p<0.001). For correct diagnosis, the agreement rate was very high at 89% [86%-91%].

**Conclusions:** Italian physicians showed a strong interest in adopting AI tools, despite significant knowledge gaps and limited practical experience. The high concordance between physicians’ evaluations and ChatGPT’s diagnostic insights suggests potential for AI-based decision support in clinical workflows. Targeted training and institutional support are essential to bridge the gap between enthusiasm and readiness for AI integration.

## Introduction

Artificial Intelligence (AI) has significantly transformed the healthcare sector, evolving from an experimental tool to an increasingly integrated technology in clinical practice. AI’s capacity to process vast amounts of data, identify complex diagnostic patterns, and support decision-making drives a paradigm shift in medical approaches, with substantial implications for healthcare quality and organisation [1]. The successful implementation of this technology relies not just on advancements but also on healthcare professionals’ acceptance and integration of AI.

While countries at the forefront of technological innovation, such as the United States and China, are allocating substantial resources to AI development and implementation [2], the challenge for other nations lies in bridging the resulting gap to benefit from these technologies and contribute to their advancement.

Italy offers a distinct perspective on healthcare AI adoption in Europe. In fact, Italy has one of the largest healthcare systems in Europe, and its investment in healthcare AI technologies is still limited compared to other EU countries. This funding gap presents significant obstacles to the widespread adoption of AI in clinical settings nationwide [3, 4].

Several pioneering AI applications have emerged in Italian healthcare institutions despite limited resources. Indeed, Italy has experienced a significant increase in AI research, particularly in medical imaging (MRI, CT, radiography), with a focus on targeting neurological diseases and cancer diagnosis. Most studies focus on machine learning for classification and segmentation tasks, showcasing an active research community [5].

At the Fondazione IRCCS Istituto Nazionale dei Tumori in Milan, an AI diagnostic imaging system has been implemented that detects early signs of lung cancer with over 90% accuracy, reducing diagnostic time by approximately 30% [6]. Similarly, the IRCCS Ospedale San Raffaele has deployed AI systems for retinal image analysis that achieve 93% sensitivity in detecting diabetic retinopathy [7]. However, these implementations remain concentrated in a few academic medical centres and have not been widely disseminated throughout the national healthcare system [8].

Despite the growing interest in AI applications in healthcare, limited empirical research has explored medical professionals’ knowledge, attitudes, and clinical agreement with AI-generated recommendations, particularly in the Italian context [9, 10].

Previous studies have highlighted a disparity in physicians’ understanding and acceptance of AI due to cultural, educational, and regulatory factors. Research on AI perception among healthcare providers has shown that, while many professionals recognise AI’s potential to enhance diagnostic and therapeutic efficiency, concerns about patient privacy, algorithmic transparency, and professional autonomy persist [11].

A systematic review of 60 studies involving over 750 physicians worldwide revealed that while more than 60% were optimistic about AI, only 15% considered it more accurate than human clinicians, with 68% believing AI should complement rather than replace physician judgment [12]. Despite this global research, a significant knowledge gap remains regarding Italian physicians’ familiarity, experience, and confidence in AI applications. A recent survey by the Italian Society of Radiology (SIRM) found that while 78% of Italian radiologists expressed interest in AI technologies, only 23% reported having received formal training, and merely 17% felt confident in their ability to critically evaluate AI-based diagnostic tools [13]. Research has identified several barriers to AI adoption in the Italian healthcare context, including insufficient training (cited by 72% of physicians), limited access to appropriate technologies (68%), concerns about data security (65%), and the absence of clear guidelines on AI implementation in clinical settings (58%). These barriers underscore the need for targeted interventions to facilitate the integration of AI into medical practice [14].

The study aims to address this gap by systematically analysing Italian physicians’ knowledge, attitudes, and level of clinical agreement with AI-generated diagnostic recommendations. The research explores physicians’ digital literacy, trust in AI technologies, and willingness to integrate them into clinical practice.

A key aspect of the research is the assessment of concordance between physicians and AI-generated diagnostic evaluations. The investigation, in fact, includes a generic clinical scenario and various speciality-specific cases, enabling an evaluation of how physicians perceive AI-suggested diagnoses and the factors influencing their level of agreement. Additionally, analysing physicians’ prior experiences with AI, their training, and perceived obstacles provides a detailed understanding of the barriers limiting technology adoption.

The research represents a crucial step in understanding Italy’s position in the global context of AI in medicine and outlining strategies to promote informed and safe adoption of these tools. The value of this study lies in its ability to provide concrete data on a highly relevant but under-explored topic in Italy, with the potential to inform both policy decisions and educational initiatives.

The research’s findings could contribute to the development of tailored interventions to overcome identified barriers and facilitate the integration of appropriate AI into clinical practice. Furthermore, the study results will help bridge the gap between Italy’s considerable medical expertise and the emerging field of healthcare AI, potentially accelerating the country’s progress in this strategically important area [3].

## Methods

### Study Design and Population

This is an observational, cross-sectional, web-based study design to investigate Italian physicians’ knowledge, attitudes, and perceptions of AI use in medicine, and to explore the level of clinical agreement between AI-generated diagnostic recommendations and those of physicians.

The study population consisted of physicians in Italy, including specialists, trainees, and general practitioners from various clinical fields and regions. Essential participation criteria included registration with the Italian Medical Council [15] at the time of the study. Exclusion criteria encompassed non-graduated or unlicensed medical students, non-physician healthcare professionals, and individuals who did not provide informed consent. The invitation to participate in the study was accompanied by a brief description of the survey, highlighting its objectives and scientific significance.

Before participating in the study, everyone received a digital informed consent form. Only after providing explicit consent could participants proceed with completing the questionnaire. The survey was deployed via LimeSurvey (v6.6.1). A comprehensive pre-launch testing phase ensured technical reliability and compatibility.

Data access was limited exclusively to authorised researchers. Participants had the right to withdraw from the study at any time.

### Questionnaire Structure

Data were collected from September 2024 to March 2025 using a structured and validated survey instrument: the I-KAPCAM-AI-Q (Knowledge, Attitudes, Practice, and Clinical Agreement between Medical Doctors and Artificial Intelligence Questionnaire) [16]. Content validity was high (CVI = 0.98) [17]. Internal consistency for the attitudinal Likert scale (8 items) was acceptable (Cronbach’s α = 0.748), while dichotomous items showed strong reliability (KR-21 = 0.832) [18, 19].

The questionnaire comprised two sections:

i. General assessment, with six domains covering demographics, AI knowledge, experience, attitudes, and willingness to use/learn about AI (including open-ended feedback).
ii. Clinical agreement, presenting real anonymised clinical scenarios submitted to ChatGPT, and evaluating physicians’ agreement with AI-generated diagnoses using a 5-point Likert scale. This section included a universal clinical scenario that all participants could respond to, as well as seven other specific scenarios tailored to participants from the following medical specialities: Hygiene and Preventive Medicine, Infectious Diseases, Obstetrics and Gynaecology, Anesthesiology, Geriatrics, Occupational Health, and Endocrinology.

The first option returned by ChatGPT in the universal scenario was the correct diagnostic option, followed by two similar options. The correct answer was presented among two other diagnoses proposed by the AI for other scenarios, which were unknown to the participants.

For specialistic scenarios, we recorded the diagnostic suggestions provided by ChatGPT in the order they appeared, without any modification, selection, or reordering by the research team. Physicians were asked to rate their level of agreement with each diagnosis on a 5-point Likert scale (1 = strongly disagree, 5 = strongly agree). Participants were not informed that the order reflected ChatGPT’s diagnostic prioritisation, to avoid influencing their evaluation.

This paper reports findings from the "universal" clinical scenario, a general case completed by all participants, describing a young adult with fatigue, fever, lymphadenopathy, and a genital lesion. Additional speciality-specific scenarios were included in the survey, but they have not been analysed in this paper. The final section collected observations and comments through two open-ended questions.

### Participant Recruitment

A non-probabilistic Snowball sampling method was adopted. This method is based on a cascade recruitment mechanism, where initial participants invite other colleagues to participate in the study, thus progressively expanding the sample. This strategy proved particularly effective in reaching many physicians operating across the national territory, belonging to different age groups and specialisations, while ensuring sample heterogeneity [20]. Various strategies were employed to achieve a broad and representative sample of the medical population, combining digital and traditional methods. First, dissemination was leveraged through social media, with particular attention to professional groups and communities on platforms such as Facebook and LinkedIn. In parallel, recruitment took place during training events, such as congresses, seminars, and refresher courses. Another method employed was word-of-mouth among colleagues, utilising participants’ professional networks to expand the study’s dissemination.

### Sample Size and Statistical Analysis

Based on a population of 439,957 Italian physicians [15], with a 5% error margin and 95% confidence interval, the estimated minimum sample size was 384 participants, calculated using the Raosoft sample size calculator [21]. However, due to the challenges of web surveys, including potential data inconsistencies and incomplete responses, we expected a dropout rate of approximately 25%. This led us to target an optimal sample size of about 500 participants to ensure adequate statistical power and representativeness even after excluding invalid responses. Web surveys present methodological challenges, including the lack of direct control over respondent identity, the possibility of unreliable responses, and the risk of selecting participants with a greater predisposition to technology, which can generate potential biases. To mitigate these risks, specific methodological measures were implemented. Completion time was controlled, excluding responses with a completion time incompatible with carefully reading the questionnaire. Data were also analysed for inconsistencies between correlated responses to identify participants who had completed the questionnaire superficially or randomly. Initially, the survey obtained 736 responses. Following quality control procedures, records with completion times significantly shorter than the estimated minimums required for thoughtful engagement with the questionnaire content were excluded. The final analysis sample consisted of 587 participants who responded to the first part of the survey and 529 who responded to the proposed scenario [22].

All variables were analysed and reported with absolute frequencies and percentages or means and standard deviations (SD) depending on the variable’s nature. Comparisons between categorical variables were made with the chi-square test, while those between continuous variables were conducted through Student’s t-test for independent samples or its non-parametric equivalent.

To assess clinical agreement between physicians and AI-generated diagnostic suggestions, we analysed physicians’ evaluations of three diagnoses proposed by ChatGPT in response to the universal clinical scenario. Descriptive statistics were used to summarise the distribution of agreement scores for each diagnosis. Agreement rates were calculated as the proportion of physicians selecting "agree" or "strongly agree" for each diagnosis, with 95% confidence intervals. The Friedman test for related samples was used to compare agreement levels across the three diagnoses. Post-hoc pairwise comparisons were performed using the Durbin-Watson test. To measure diagnostic discrimination, we identified physicians who both strongly agreed with the correct diagnosis (score ≥4) and rejected incorrect diagnoses. Statistical significance was assessed using an alpha level of 0.05. All analyses were conducted with Stata 18 software.

## Results

The demographic characteristics of participants revealed a relatively balanced gender distribution with a slight female predominance (57.2% female, 42.4% male). The mean age was 42.7 years (SD = 13.6), with participants reporting an average of 14.1 years of clinical practice experience (SD = 13.2). Geographical distribution showed a concentration in Central Italy (58.8%), followed by Southern Italy and the Islands (28.6%), and Northern Italy (12.6%). Regarding professional status, 60.5% were specialist physicians and 39.5% were residents. The most represented medical specialisations were Hygiene and Preventive Medicine (16.0%), Gynaecology and Obstetrics (11.4%), and Anesthesiology and Intensive Care (10.6%). Other notable specialities included Geriatrics (7.3%), Occupational Medicine (7.5%), Endocrinology and Metabolic Diseases (6.1%) and Infectious and Tropical Diseases (6.1%). The remaining 35.0% represented various other specialisations. Only 18.4% of participants reported receiving specific training in digital or information technologies during their education, highlighting a substantial educational deficit in this rapidly evolving field.

Table 2 shows that most physicians (64.8%) have only basic knowledge, while 21.6% have no knowledge at all, according to their self-assessment of AI knowledge. Concerning familiarity with specific AI applications, diagnostic imaging was the most recognised (47.2%), likely reflecting the earlier adoption and more widespread implementation of AI in radiology and dermatology. Other relatively familiar applications included report interpretation (33.2%), analysis of electronic health records (27.9%), and clinical decision support systems (25.4%).

**Table 1.** Demographic Characteristics of Study Participants.

|  |  |
| --- | --- |
| Characteristic | N=587 |
| Gender | N (%) |
| - Male | 249 (42.4%) |
| - Female | 336 (57.2%) |
| - Other | 2 (0.3%) |
| Age (years) | 42.726 (13.641) |
| Geographic area | N (%) |
| - North | 74 (12.6%) |
| - Central | 345 (58.8%) |
| - South and Islands | 168 (28.6%) |
| Medical specialisation | N (%) |
| - Geriatrics | 43 (7.3%) |
| - Endocrinology and Metabolic Diseases | 36 (6.1%) |
| - Infectious and Tropical Diseases | 36 (6.1%) |
| - Gynaecology and Obstetrics | 67 (11.4%) |
| - Anesthesiology and Intensive Care | 62 (10.6%) |
| - Hygiene and Preventive Medicine | 94 (16.0%) |
| - Occupational Medicine | 44 (7.5%) |
| - Other specialisations | 205 (35.0%) |
| Professional status | N (%) |
| - Specialist physician | 355 (60.5%) |
| - Resident | 232 (39.5%) |
| Years of practice | 14.109 (13.221) |
| Specific training in digital or information technologies | N (%) |
| - No | 479 (81.6%) |
| - Yes | 108 (18.4%) |

*Note: Data are presented as N (%) for categorical variables and mean (SD) for continuous variables.*

**Table 2.** Knowledge and Familiarity with AI in Medicine (N=587).

| Variable | N (%) |
| --- | --- |
| Self-assessed knowledge of artificial intelligence |  |
| No knowledge | 127 (21.6%) |
| Basic knowledge | 380 (64.8%) |
| Intermediate knowledge | 67 (11.4%) |
| Advanced knowledge | 13 (2.2%) |
| Familiarity with AI applications in medicine |  |
| Diagnostic imaging (radiology, dermatology) | 277 (47.2%) |
| Clinical decision support systems | 149 (25.4%) |
| Care planning and management | 63 (10.7%) |
| Patient data monitoring and analysis | 147 (25.0%) |
| Report interpretation (ECG, EEG, etc.) | 195 (33.2%) |
| Machine learning in diagnosis | 95 (16.2%) |
| Machine learning and Big Data for new drug development | 56 (9.5%) |
| Virtual nursing assistants | 10 (1.7%) |
| Chatbots for diagnostic queries | 136 (23.2%) |
| Monitoring, home care and predictive analysis | 44 (7.5%) |
| Personalised medicine | 50 (8.5%) |
| Improving genomic editing | 16 (2.7%) |
| Electronic health records and data analysis | 164 (27.9%) |
| AI-based robotic surgery procedures | 65 (11.1%) |
| None of the above | 162 (27.6%) |

The use of artificial intelligence instruments in clinical practice throughout their life was stated by 21.6% (127) of all interviewees. Analysis of AI adoption (instruments or applications) among medical professionals in the last year reveals its limited integration into clinical practice. Diagnostic applications exhibit the highest usage rates, with diagnostic imaging (7.7%), diagnostic chatbots (7.2%), and clinical decision support systems (6.3%) leading the way. Moderate adoption exists in report interpretation (4.8%), Electronic health records (EHR) data analysis (4.3%), and patient monitoring (3.9%). Most advanced AI applications demonstrate minimal penetration: machine learning for diagnosis (2.6%), personalised medicine (1.5%), home care monitoring (1.2%), robotic surgery (0.5%), and genomic/pharmaceutical applications (0.2%) (Table 3).

**Table 3.** AI Tools (instruments or applications) Usage Among Medical Professionals in the last year before the interview.

| **AI Tool/Application** |  |  |
| --- | --- | --- |
| Use of artificial intelligence instruments in clinical practice | 127/587 (21.6%) |  |
|  | **Usage (Yes)**<br>**N=587** | **Usage (Yes)**<br>**N=127** |
| Diagnostic imaging (radiology, dermatology) | 45/587 (7.7%) | 45/127 (35.4%) |
| Clinical decision support systems | 37/587 (6.3%) | 37/127 (29.1%) |
| Care planning and management | 15/587 (2.6%) | 15/127 (11.8%) |
| Patient data monitoring and analysis | 23/587 (3.9%) | 23/127 (18.1%) |
| Report interpretation (ECG, EEG, etc.) | 28/587 (4.8%) | 28/127 (22.0%) |
| Machine learning in diagnosis | 15/587 (2.6%) | 15/127 (11.8%) |
| Drug discovery and development | 1/587 (0.2%) | 1/127 (0.8%) |
| Virtual nursing assistants | 1/587 (0.2%) | 1/127 (0.8%) |
| Diagnostic chatbots | 42/587 (7.2%) | 42/127 (33.1%) |
| Home care monitoring and predictive analysis | 7/587 (1.2%) | 7/127 (5.5%) |
| Personalised medicine | 9/587 (1.5%) | 9/127 (7.1%) |
| Genomic editing enhancing | 1/587 (0.2%) | 1/127 (0.8%) |
| Electronic health records (EHR) and data analysis | 25/587 (4.3%) | 25/127 (19.7%) |
| AI-based robotic surgery procedures | 3/587 (0.5%) | 3/127 (2.4%) |

As reported in Table 4, analysis of the Italian Physicians’ attitudes toward Artificial Intelligence revealed predominantly positive attitudes toward AI among Italian physicians. The highest agreement was observed regarding the need for specific AI training (92.3% agreed/strongly agreed, mean=4.29±0.72). Strong positive perceptions were also found in AI’s potential to aid in differential diagnosis (77.7% agreed/strongly agreed, mean=3.84±0.66) and improve prescription of diagnostic tests (68.8% agreed/strongly agreed, mean=3.68±0.74). Physicians showed moderate optimism about AI enhancing workflow efficiency, with 65.4% believing AI could improve patient management (mean=3.61±0.85) and 61.8% acknowledging AI’s potential to reduce workload (mean=3.63±0.91). Participants did not perceive AI as a threat to their professional role (56.4% disagreed/strongly disagreed, mean=2.45±0.95), 31.5% expressed neutrality.

**Table 4.** Italian Physicians’ Attitudes Toward Artificial Intelligence in Medicine n=587.

|  | Strongly Disagree | Disagree | Neutral | Agree | Strongly Agree |  |  |
| --- | --- | --- | --- | --- | --- | --- | --- |
| Statement | n (%) | n (%) | n (%) | n (%) | n (%) | Median [IQR] | Mean ± SD |
| AI can aid in differential diagnosis | 4 (0.68) | 16 (2.73) | 111 (18.91) | 394 (67.12) | 62 (10.56) | 4 [4-4] | 3.84 ± 0.66 |
| AI can improve therapeutic prescriptions | 7 (1.32) | 42 (7.94) | 132 (24.95) | 302 (57.09) | 46 (8.70) | 4 [3-4] | 3.64 ± 0.80 |
| AI can improve the prescription of diagnostic and laboratory tests | 8 (1.36) | 31 (5.28) | 144 (24.53) | 364 (62.01) | 40 (6.81) | 4 [3-4] | 3.68 ± 0.74 |
| AI can reduce physicians' workload | 9 (1.53) | 63 (10.73) | 152 (25.89) | 279 (47.53) | 84 (14.31) | 4 [3-4] | 3.63 ± 0.91 |
| AI can improve efficiency in patient management | 12 (2.04) | 53 (9.03) | 138 (23.51) | 331 (56.39) | 53 (9.03) | 4 [3-4] | 3.61 ± 0.85 |
| AI represents a threat to the role of physicians | 85 (14.48) | 246 (41.91) | 185 (31.52) | 52 (8.86) | 19 (3.24) | 2 [2-3] | 2.45 ± 0.95 |
| Using AI requires specific additional training for physicians | 6 (1.02) | 7 (1.19) | 32 (5.45) | 306 (52.13) | 236 (40.20) | 4 [4-5] | 4.29 ± 0.72 |

As reported in Table 5, the most striking finding is the prominent role of education and training. "Lack of training" is identified as the primary barrier by 76.7% of physicians, while "professional training and education" is overwhelmingly selected as the top incentive (85.7%). There was an alignment between identified barriers and corresponding incentives: the barrier of "lack of scientific evidence" (27.6%) corresponds with the incentive of "scientific evidence on efficacy and safety" (55.2%); "the cost of AI technologies" (42.9%) pairs with "the cost reduction" (28.8%), "the resistance to change" (50.9%) may be addressed through "the continuous technical support" (52.3%).

**Table 5.**
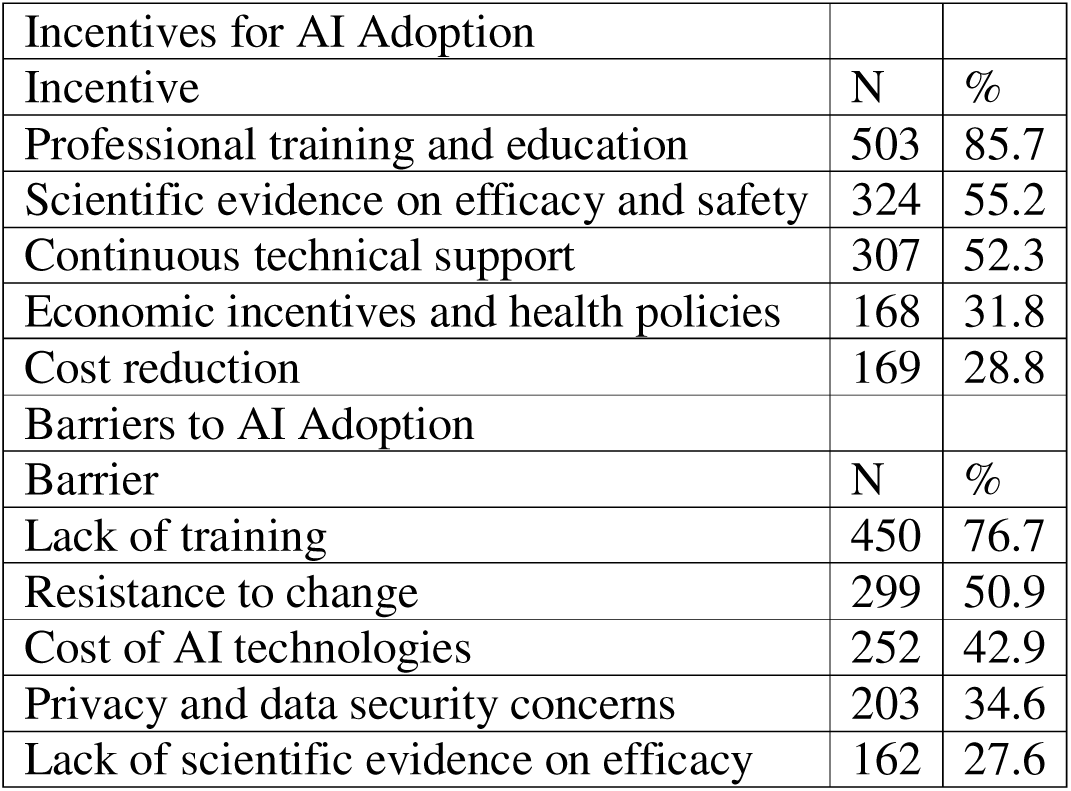
Incentives and Barriers to AI Adoption Among Italian Physicians.

| Incentives for AI Adoption |  |  |
| --- | --- | --- |
| Incentive | N | % |
| Professional training and education | 503 | 85.7 |
| Scientific evidence on efficacy and safety | 324 | 55.2 |
| Continuous technical support | 307 | 52.3 |
| Economic incentives and health policies | 168 | 31.8 |
| Cost reduction | 169 | 28.8 |
| Barriers to AI Adoption |  |  |
| Barrier | N | % |
| Lack of training | 450 | 76.7 |
| Resistance to change | 299 | 50.9 |
| Cost of AI technologies | 252 | 42.9 |
| Privacy and data security concerns | 203 | 34.6 |
| Lack of scientific evidence on efficacy | 162 | 27.6 |

The analysis reveals a striking readiness for AI adoption, with 81.9% of physicians reporting willingness to integrate AI into their clinical practice. Consistently, most physicians (93.5%) expressed interest in receiving AI training, with a preference for online courses (64.7%) and practical hands-on sessions (61.2%), followed by in-person seminars (47.6%) and webinars (29.0%). (Table 6).

**Table 6.** Willingness and Training Preferences (n=587).

| Category | Subcategory | N | Percentage (%) |
| --- | --- | --- | --- |
| Willingness to Integrate AI | Very willing | 153 | 26.0 |
|  | Willing | 328 | 55.9 |
|  | Undecided | 78 | 13.3 |
|  | Somewhat unwilling | 20 | 3.4 |
|  | Not at all willing | 8 | 1.4 |
| Interest in AI Training | Yes | 549 | 93.5 |
|  | No | 38 | 6.5 |
| Preferred Training Modalities | Online courses | 357 | 64.7 |
|  | Practical hands-on training | 339 | 61.2 |
|  | In-person seminars and workshops | 263 | 47.6 |
|  | Webinars and conferences | 160 | 29.0 |

### Clinical agreement between ChatGPT and Physicians

To assess physician agreement with ChatGPT’s diagnostic suggestions, Diagnosis_a was identified as the correct diagnosis, followed by Diagnosis_b and Diagnosis_c. The analysis reveals a clear hierarchy in physicians’ level of agreement with the three diagnoses proposed by ChatGPT. Physicians show significantly higher agreement with Diagnosis_a (mean = 4.07, median = 4), which was the correct diagnosis and was proposed as the first option by ChatGPT. This is followed by lower agreement with Diagnosis_c (mean = 2.57, median = 2), while showing the lowest agreement with Diagnosis_b (mean = 1.82, median = 2). The Friedman test demonstrates significant differences in agreement levels among the three diagnoses (χ² = 735, df = 2, p < 0.001). All pairwise comparisons using the Durbin-Conover test show statistically significant differences (all p < 0.001), confirming that physicians’ agreement levels differ significantly across all three diagnoses. The physician responses for Diagnosis_a (the correct diagnosis) display the lowest variability (SD = 0.75, IQR = 0), which indicates strong consensus among medical professionals in favour of the correct diagnosis. Diagnosis_c’s reactions have the most variability (SD = 1.05, IQR = 1) (Table 7, Figure 2).

**Figure 2.**
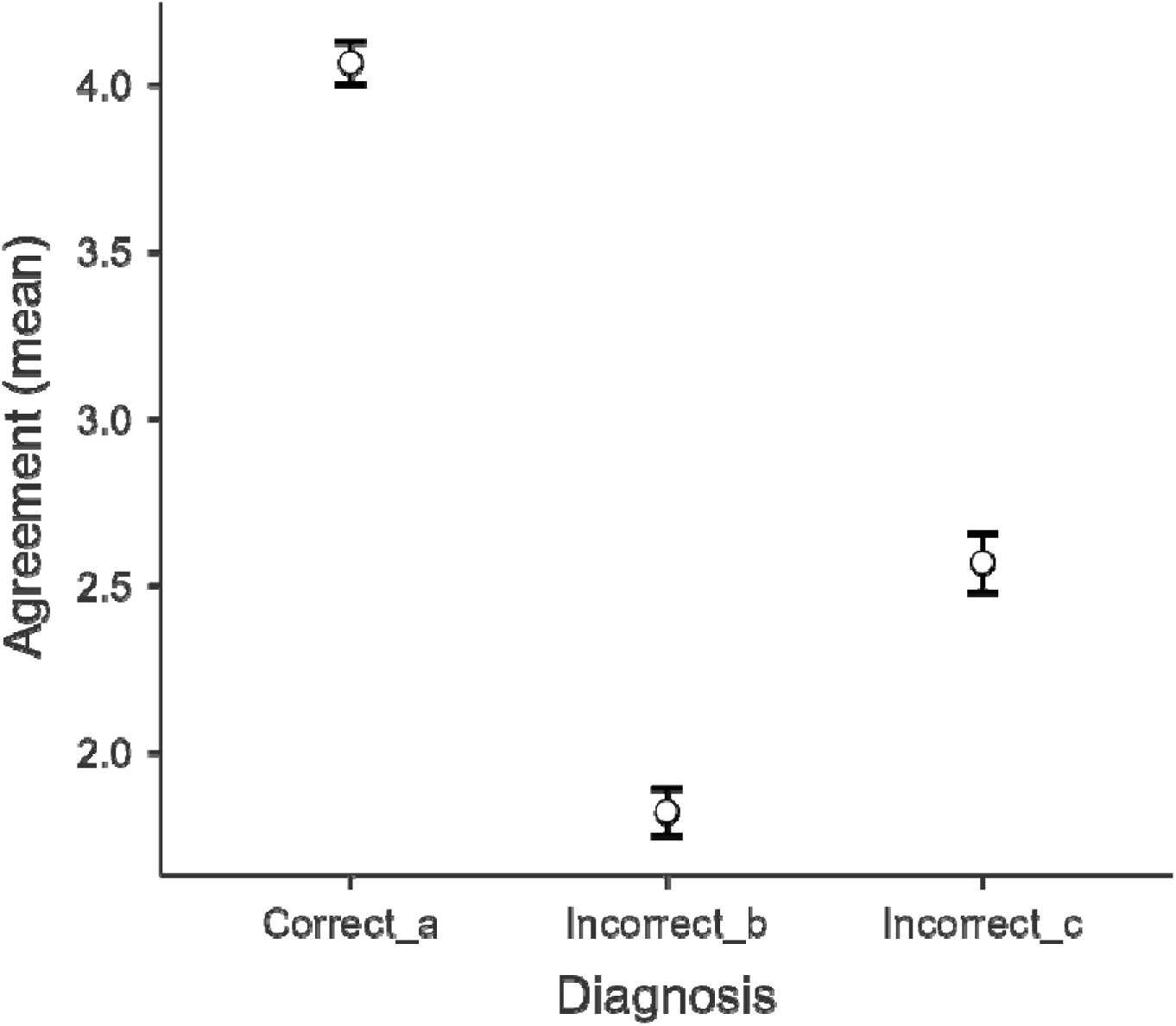
Clinical Agreement Distribution.

**Table 7.** Clinical agreement analysis between physicians and AI diagnoses for the universal scenario only.

|  | Diagnosis_a | Diagnosis_b | Diagnosis_c |
| --- | --- | --- | --- |
| Descriptive Statistics | N=529 | N=529 | N=529 |
| Mean (SD) | 4.07 (0.74) | 1.82 (0.81) | 2.57 (1.05) |
| Median (IQR) | 4 (0) | 2 (1) | 2 (1) |
| Friedman Test | $\chi^2 = 735$, $df = 2$, $p < 0.001$ | | |
| Pairwise Comparisons | Statistic | p-value |  |
| Diagnosis_a - Diagnosis_b | 48.6 | < 0.001 |  |
| Diagnosis_a- Diagnosis_c | 29.5 | < 0.001 |  |
| Diagnosis_b- Diagnosis_c | 19.1 | < 0.001 |  |

A deeper analysis of concordance patterns reveals that for Diagnosis_a, the agreement rate (agree or strongly agree) was 89% [95%CI: 86%-92%]. For Diagnosis_b, the agreement rate was very low at 4% [95%CI: 2%-6%], while the agreement rate was 23% [95%CI: 20%-27%] for Diagnosis_c. 68% [95%CI: 64%-72%] of physicians demonstrated clear diagnostic discrimination, both strongly agreeing with the correct diagnosis (score ≥4).

## Discussion

This national cross-sectional web-based study investigated the knowledge, attitudes, and clinical concordance with AI-generated diagnostic recommendations among 587 Italian physicians, including both residents and trained medical doctors, conducted from September 2024 to March 2025. Our findings reveal significant knowledge gaps, strong interest in AI adoption, limited current usage patterns, and remarkably high clinical agreement between physicians and ChatGPT’s diagnostic recommendations.

The following results provide important insights into AI integration’s current state and future potential in Italian healthcare.

### Knowledge Gap and Educational Needs

Significant gaps in AI knowledge and training among Italian physicians were found. Only 18.4% of participants reported receiving specific training in digital or information technologies during their education, highlighting a substantial educational deficit in this rapidly evolving field. This aligns with findings from Blease et al. [23] and Sapci [24], who identified similar patterns of limited AI education across European healthcare systems. In a very recent synthesis of literature and case studies [25], the dual nature of AI—its promising benefits juxtaposed with significant challenges concerning privacy, security, autonomy, and freedom- is highlighted. Authors underscore the vital importance of public acceptance, normative frameworks, technological innovation, and international collaboration in effectively addressing these issues. Most physicians (64.8%) self-assessed as possessing only basic knowledge of AI, with 2.2% claiming advanced knowledge. This knowledge deficiency creates a barrier to adoption that needs to be addressed through targeted educational interventions. Similar knowledge gaps have been documented in other European countries, suggesting a systemic challenge in medical education systems that have not yet fully incorporated digital health competencies into their curricula [24, 26].

### Familiarity with AI Applications

When examining familiarity with specific AI applications, diagnostic imaging emerged as the most recognised (47.2%), likely reflecting the earlier adoption and more widespread implementation of AI in radiology and dermatology [27]. This finding is consistent with the literature, as imaging specialities have pioneered AI integration due to the structured nature of their data and clear pattern recognition tasks [1, 28]. Other relatively familiar applications included report interpretation (33.2%), electronic health records analysis (27.9%), and clinical decision support systems (25.4%). Despite the growing prominence of AI in healthcare, a substantial proportion (27.6%) of physicians reported no familiarity with any of the listed AI applications. Applications with the lowest recognition included virtual nursing assistants (1.7%), genomic editing (2.7%), and home care monitoring systems (7.5%), indicating areas where awareness campaigns might be particularly beneficial. This distribution of familiarity mirrors findings from Blease et al. [23], who documented similar patterns of awareness among primary care physicians across Europe and North America.

### AI Adoption Patterns

The integration of AI into clinical practice was limited, with only 127 (22%) of respondents reporting the use of AI tools. The analysis revealed notable patterns, particularly the significant uptake of AI chatbots. Diagnostic chatbots [42/127 (33.1%)] represented one of the most widely adopted AI applications, nearly matching diagnostic imaging tools [45/127 (35.4%)] and surpassing most other AI technologies. This growing trend in chatbot utilisation aligns with recent systematic reviews that have documented the increasing implementation of conversational AI in clinical settings [29].

Clinical decision support systems [37/127 (29.1%)] follow closely, while other applications show moderate to minimal adoption, including report interpretation, EHR data analysis, and patient monitoring. The substantial adoption of chatbots relative to other technologies suggests that text-based, conversational AI interfaces may offer advantages in clinical workflows, potentially due to their accessibility, lower implementation barriers, and alignment with traditional consultation processes [30]. Laymouna et al. [31] similarly observed that chatbots often serve as "gateway technologies" for broader healthcare AI adoption due to their intuitive interfaces and integration with existing clinical communication patterns.

### Attitudes Toward AI

Italian physicians recognise AI’s potential value in clinical decision support while acknowledging the importance of proper training. The highest agreement was observed regarding the need for specific AI training (92.3% agreed/strongly agreed, mean=4.29±0.71), indicating widespread recognition that effective AI implementation requires dedicated education. This finding aligns with multinational surveys conducted by Pinto et al. [32], which documented similar educational priorities among European physicians. Most respondents did not perceive AI as a threat to their professional role (56.4% disagreed/strongly disagreed, mean=2.45±0.95), though 31.5% expressed neutrality on this issue, suggesting some uncertainty about long-term implications. Positive attitudes towards AI coexist with some concerns about its impact on the workforce. This indicates a practical approach to adopting AI, which can help integrate these technologies thoughtfully into healthcare systems [32, 34].

### Barriers and Incentives for AI Adoption

The parallel structure between identified barriers and incentives suggests that physicians clearly understand both the challenges and potential solutions for AI integration. The high percentage of physicians citing "resistance to change" (50.9%) reflects the well-documented conservatism in medical practice, where practitioners are understandably cautious about adopting new technologies that might affect patient care. This finding is consistent with Jussupow et al. [35], who identified professional identity threats as a significant factor in resistance to AI adoption among medical professionals.

However, the equally high percentage seeking "continuous technical support" (52.3%) shows a willingness to overcome this resistance with proper assistance. Similar findings by Castagno et al. [36] emphasised the critical importance of ongoing technical support in successful healthcare AI implementations.

Privacy and data security concerns (34.6%) represent a moderate barrier, reflecting the growing awareness of the ethical and legal implications of AI in healthcare. This is particularly relevant in the European context, where strict GDPR regulations apply. Gille et al. [37] documented similar privacy concerns among healthcare providers in GDPR-regulated environments, highlighting the need for transparent AI governance frameworks.

While the cost of AI technologies is cited as a significant barrier (42.9%), economic incentives (31.8%) and cost reduction (28.8%) rank lowest among potential incentives. This suggests that Italian physicians are more motivated by professional improvement and evidence-based practice than by financial considerations, a pattern also observed by Liyanage et al. [38] in their analysis of Physician Technology Adoption Drivers. Professional training and education (85.7%) emerged as the strongest incentive, together with scientific evidence on AI efficacy and safety (55.2%), underscoring physicians’ preference for knowledge-based and evidence-driven support over purely economic motivations

### Readiness for AI Integration

Data results reveal a striking readiness for AI adoption, with 81.9% of physicians reporting willingness to integrate AI into their clinical practice. This openness is particularly noteworthy given the identified barriers, primarily lack of training (76.7%) and resistance to change (50.9%). The findings highlight a critical knowledge-action gap: while physicians recognise AI’s potential, they clearly identify education as both the primary barrier and solution. An overwhelming 93.5% express interest in AI training programs, with a preference for accessible formats, such as online courses (64.7%) and hands-on training (61.2%).

These results suggest that Italian healthcare is poised for AI transformation, provided that appropriate educational infrastructure and evidence-based implementation strategies are prioritised. Similar patterns of readiness coupled with educational needs have been documented by Huisman et al. [39] across multiple European healthcare systems.

### AI-Physician Diagnostic Concordance

The analysis shows a significant alignment between physicians’ consensus and ChatGPT’s diagnostic prioritisation. The clear hierarchy in agreement levels (4.06 for the correct diagnosis versus 2.57 and 1.82 for incorrect alternatives) suggests that ChatGPT successfully captures expert clinicians’ differential diagnostic process. The statistical significance of the differences between all three diagnoses (p<0.001) further confirms that physicians’ evaluations clearly distinguished between the diagnostic options in a way that aligns with ChatGPT’s proposal [89%, 95% CI: 86%- 91%].

These findings align with a recent work by Nori et al. [40], who documented similar levels of concordance between LLM diagnostic outputs and physician judgement.

This concordance between physician judgement and AI-generated diagnostic recommendations has substantial implications for clinical practice and AI implementation in healthcare. It suggests that Large Language Models like ChatGPT demonstrate the ability to emulate human diagnostic reasoning processes, despite not being trained explicitly for medical diagnoses. From a practical perspective, this concordance opens concrete possibilities for integrating AI as decision support in daily clinical practice. In settings characterised by high workloads, such as emergency departments or primary care, LLM-based tools could serve as an automated "second opinion," validating the physician’s diagnostic orientation or suggesting alternative diagnoses worthy of consideration. Thirunavukarasu et al. [41] similarly concluded that general-purpose LLMs show promise as clinical decision support tools, particularly in high-volume clinical environments.

The ability of ChatGPT to propose diagnoses aligned with collective medical judgment could be particularly valuable for less experienced clinicians, including physicians in training or professionals working outside their specialisation area. AI could thus represent a tool to support continuing education, and a mechanism to reduce variability in clinical decisions, as suggested by Magrabi et al. [42] in their analysis of AI as an educational adjunct in medical training. However, the implications of this concordance should also be considered in terms of potential risks.

Excessive confidence in the alignment between AI and medical judgment could lead to "collective blindness" in atypical or rare cases, where both AI and physicians might share the same biases or knowledge limitations. Paradoxically, cases where ChatGPT showed less concordance with physicians might represent the most interesting areas for mutual learning and improvement of diagnostic systems. Daneshjou et al. [43] highlighted similar concerns regarding potential reinforcement of existing biases when AI and human judgment are overly concordant.

From a regulatory and organisational perspective, this significant concordance provides empirical evidence supporting the implementation of AI systems as augmentation tools, rather than replacements for clinical judgment. The fact that physicians and AI converge on similar diagnoses suggests the possibility of developing integrated clinical workflows where AI plays a complementary role, validating and enriching the medical decision-making process.

### Strengths and Limitations

This study presents several strengths, including the combined assessment of knowledge, attitudes, and clinical concordance in one investigation, which provides a holistic view of AI integration challenges. The innovative methodology of using AI-generated diagnoses and measuring physician concordance provides concrete data on the real-world utility of AI, an approach that addresses the gap between theoretical AI capabilities and practical clinical applications highlighted by Sendak et al. [44]. However, several limitations should be considered when interpreting these results. First, the convenience sampling method, the snowball technique, does not guarantee the sample’s representativeness compared to the entire Italian physician population. Consequently, the results cannot be generalised to the national population with full reliability. Additionally, there may have been a selection bias related to a greater propensity to participate among subjects interested in artificial intelligence.

A methodological consideration concerns the online survey format and the self-reported nature of the data. Because the survey was conducted remotely, participants theoretically could have consulted various resources, including ChatGPT, when responding to the clinical scenarios. To mitigate this risk, we implemented specific quality control measures, including carefully monitoring completion times. Responses with completion times incompatible with thoughtful engagement (either too short, suggesting random answers, or excessively long, suggesting external consultation) were excluded from the analysis. This filtering process substantially reduced the likelihood that participants consulted AI tools during the completion process, strengthening the validity of our concordance findings. Nevertheless, we cannot eliminate the possibility that some participants may have used external resources, which represents a limitation inherent to remote survey methodologies. Another limitation relates to presenting a universal clinical scenario for assessing clinical concordance. While this approach ensured comparability across all participants, it may not capture the complexity and diversity of real-world clinical decision-making across different medical specialities and patient presentations. The findings are limited to the case presented and may not generalise to other clinical contexts or more uncertain diagnostic situations.

Despite these limitations, the sample size achieved exceeds that calculated as necessary to ensure acceptable precision in descriptive estimates, and the quality control procedures implemented enhance the reliability of the collected data. The exploratory analysis has highlighted useful trends and perceptions to guide more extensive future studies with rigorous designs.

## Conclusions

Our findings provide an empirical foundation to support the evidence-based integration of AI tools in Italian clinical workflows, while highlighting the urgent need for structured education programs, tailored to medical professionals. Italian physicians show a strong interest in adopting AI tools, despite significant knowledge gaps and limited practical experience. The high concordance between physicians’ evaluations and ChatGPT’s diagnostic proposals suggests potential for AI-based decision support in clinical workflows. Targeted training and institutional support are essential to bridge the gap between enthusiasm and readiness for AI integration.

The study reveals that Italian physicians are cautiously optimistic about AI adoption, with a strong emphasis on proper training, scientific validation, and technical support as prerequisites for successful implementation in clinical practice. The remarkable alignment between physician judgment and AI-generated diagnostic recommendations opens new possibilities for integrating AI as decision support tools in clinical practice, provided that appropriate educational infrastructure and evidence-based implementation strategies are prioritised.

## Data Availability

The raw data supporting the conclusions of this article will be made available by the authors
upon request.

## Statements and Declarations

### Author Contributions

All authors contributed to the study conception and design. Material preparation, data collection and analysis were performed by Vincenza Cofini, Eugenio Benvenuti, Mario Muselli and Stefano Necozione. The first draft of the manuscript was written by Vincenza Cofini, Eugenio Benvenuti, Laura Piccardi, Martina Mancinelli, Mario Muselli, and all authors commented on previous versions of the manuscript. All authors read and approved the final manuscript.

### Data Availability

The datasets generated during and/or analysed during the current study are available from the corresponding author on reasonable request.

## References

1. Esteva A, Robicquet A, Ramsundar B, Kuleshov V, DePristo M, Chou K, Cui C, Corrado G, Thrun S, Dean J. A guide to deep learning in healthcare. Nat Med. 2019 Jan;25(1):24–29. doi: 10.1038/s41591-018-0316-z

2. Maslej N, Fattorini L, Perrault R, Parli V, Reuel A, Brynjolfsson E, Etchemendy J, Ligett K, Lyons T, Manyika J, Niebles JC, Shoham Y, Wald R, Clark J. The AI Index 2024 Annual Report. AI Index Steering Committee, Institute for Human-Centered AI, Stanford University, 2024, Stanford, CA.

3. Ministero della Salute. (2023). Piano nazionale per l’innovazione del sistema sanitario basata sull’intelligenza artificiale 2023-2027. Roma: Ministero della Salute.

4. Cascini F, Beccia F, Causio FA, Melnyk A, Zaino A, Ricciardi W. Scoping review of the current landscape of AI-based applications in clinical trials. Front Public Health. 2022 Aug 12;10:949377. doi: 10.3389/fpubh.2022.949377

5. Reale R, Biasin E, Scardovi A, Toro S. The Design and Implementation of a National AI Platform for Public Healthcare in Italy: Implications for Semantics and Interoperability. ArXiv, abs/2304.11893. 2023. 10.48550/arXiv.2304.11893

6. Cellina M, Cacioppa LM, Cè M, Chiarpenello V, Costa M, Vincenzo Z, Pais D, Bausano MV, Rossini N, Bruno A, Floridi C. Artificial Intelligence in Lung Cancer Screening: The Future Is Now. Cancers (Basel). 2023 Aug 30;15(17):4344. doi: 10.3390/cancers15174344

7. Injante R, Julca M. Detection of diabetic retinopathy using artificial intelligence: an exploratory systematic review. LatIA. 2024; 2:112. 10.62486/latia2024112

8. Ahmadi A. Navigating the Future: Challenges and Opportunities in Hospital Care in Italy - A Review of AI and Big Data Integration. International Journal of BioMed Insights. 2024,1(1), 23-34. doi: 10.22034/ijbmi.2024.198709.

9. Bini, S. A. (2021). Artificial intelligence, machine learning, deep learning, and cognitive computing: What do these terms mean and how will they impact health care? Journal of Arthroplasty, 33(8), 2358–2361. 10.1016/j.arth.2018.02.067

10. Cingolani, M., Scendoni, R., Fedeli, P., & Cembrani, F. (2023). Artificial intelligence and digital medicine for integrated home care services in Italy: Opportunities and limits. Frontiers in public health, 10, 1095001. 10.3389/fpubh.2022.1095001.

11. Commissione europea, D. G. (2019). Orientamenti etici per un’IA affidabile. https://data.europa.eu/doi/10.2759/640340

12. Chen M, Zhang B, Cai Z, Seery S, Gonzalez MJ, Ali NM, Ren R, Qiao Y, Xue P, Jiang Y. Acceptance of clinical artificial intelligence among physicians and medical students: A systematic review with cross-sectional survey. Front Med (Lausanne). 2022 Aug 31;9:990604. doi: 10.3389/fmed.2022.990604

13. Neri E, Coppola F, Miele V, Bibbolino C, Grassi R. Artificial intelligence: Who is responsible for the diagnosis? Radiol Med. 2020 Jun;125(6):517–521. doi: 10.1007/s11547-020-01135-9. Epub 2020 Jan 31.

14. Associazione Italiana di Informatica Medica. Indagine nazionale sull’adozione dell’intelligenza artificiale nella pratica clinica in Italia. Milano: AIIM Press; 2023.

15. FNOMCeO. Medici e Odontoiatri: ecco i numeri della professione. 2023. https://portale.fnomceo.it/medici-e-odontoiatri-ecco-i-numeri-della-professione/. Accessed 24 Oct 2025.

16. Cofini V, Piccardi L, Benvenuti E, Di Pangrazio G, Cimino E, Mancinelli M, Muselli M, Petrucci E, Picchi G, Palermo P, Tobia L, Barbonetti A, Desideri G, Guido M, Marinangeli F, Fabiani L, Necozione S. The I-KAPCAM-AI-Q: a novel instrument for evaluating health care providers’ AI awareness in Italy. Front Public Health. 2025 Sep 18;13:1655659. doi: 10.3389/fpubh.2025.1655659

17. Dalawi I, Isa MR, Chen XW, Azhar ZI, Aimran N. Development of the Malay Language of understanding, attitude, practice and health literacy questionnaire on COVID-19 (MUAPHQ C-19): content validity & face validity analysis. BMC Public Health. 2023 Jun 13;23(1):1131. doi: 10.1186/s12889-023-16044-5.

18. Bland JM, Altman DG. Statistics notes: Cronbach’s alpha. BMJ. 1997. 314(7080), 572. 10.1136/bmj.314.7080.572

19. Foster RC. KR20 and KR21 for Some Nondichotomous Data (It’s Not Just Cronbach’s Alpha). Educational and Psychological Measurement. 2021. 81(6), 1172–1185. 10.1177/0013164421992535

20. Sadler GR, Lee HC, Lim RS, Fullerton J. Recruitment of hard-to-reach population subgroups via adaptations of the snowball sampling strategy. Nurs Health Sci. 2010 Sep 1;12(3):369–74. doi: 10.1111/j.1442-2018.2010.00541.x

21. Raosoft Inc. Sample size calculator. 2024. http://www.raosoft.com/samplesize.html. Accessed 24 Oct 2025.

22. Cofini V, Benvenuti E, Mancinelli M, Di Pangrazio G, Piccardi L, Muselli M, et al. A Framework to Improve Data Quality and Manage Dropout in Web-Based Medical Surveys: Insights from an Ai Awareness Study among Italian Physicians. ebph [Internet]. 2025. https://riviste.unimi.it/index.php/ebph/article/view/29202. Accessed 24 Oct 2025.

23. Blease C, Kharko A, Bernstein M, Bradley C, Houston M, Walsh I, Hägglund M, DesRoches C, Mandl KD. Machine learning in medical education: a survey of the experiences and opinions of medical students in Ireland. BMJ Health Care Inform. 2022 Feb;29(1):e100480. doi: 10.1136/bmjhci-2021-100480

24. Sapci AH, Sapci HA. Artificial Intelligence Education and Tools for Medical and Health Informatics Students: Systematic Review. JMIR Med Educ. 2020 Jun 30;6(1):e19285. doi: 10.2196/19285

25. Ding X, Shang B, Xie C, Xin J, Yu F. Artificial intelligence in the COVID-19 pandemic: balancing benefits and ethical challenges in China’s response. Humanit Soc Sci Commun. 2025. 12, 245. 10.1057/s41599-025-04564-x

26. Wartman SA, Combs CD. Medical Education Must Move From the Information Age to the Age of Artificial Intelligence. Acad Med. 2018 Aug;93(8):1107–1109. doi: 10.1097/ACM.0000000000002044

27. Pesapane F, Codari M, Sardanelli F. Artificial intelligence in medical imaging: threat or opportunity? Radiologists again at the forefront of innovation in medicine. Eur Radiol Exp. 2018 Oct 24;2(1):35. doi: 10.1186/s41747-018-0061-6

28. Hosny A, Parmar C, Quackenbush J, Schwartz LH, Aerts HJWL. Artificial intelligence in radiology. Nat Rev Cancer. 2018 Aug;18(8):500–510. doi: 10.1038/s41568-018-0016-5

29. Tudor Car L, Dhinagaran DA, Kyaw BM, Kowatsch T, Joty S, Theng YL, Atun R. Conversational Agents in Health Care: Scoping Review and Conceptual Analysis. J Med Internet Res. 2020 Aug 7;22(8):e17158. doi: 10.2196/17158

30. Ma Y, Achiche S, Pomey MP, Paquette J, Adjtoutah N, Vicente S, Engler K; MARVIN chatbots Patient Expert Committee; Laymouna M, Lessard D, Lemire B, Asselah J, Therrien R, Osmanlliu E, Zawati MH, Joly Y, Lebouché B. Adapting and Evaluating an AI-Based Chatbot Through Patient and Stakeholder Engagement to Provide Information for Different Health Conditions: Master Protocol for an Adaptive Platform Trial (the MARVIN Chatbots Study). JMIR Res Protoc. 2024 Feb 13;13:e54668. doi: 10.2196/54668

31. Laymouna M, Ma Y, Lessard D, Schuster T, Engler K, Lebouché B. Roles, Users, Benefits, and Limitations of Chatbots in Health Care: Rapid Review. J Med Internet Res. 2024 Jul 23;26:e56930. doi: 10.2196/56930

32. Pinto Dos Santos D, Giese D, Brodehl S, Chon SH, Staab W, Kleinert R, Maintz D, Baeßler B. Medical students’ attitude towards artificial intelligence: a multicentre survey. Eur Radiol. 2019 Apr;29(4):1640–1646. doi: 10.1007/s00330-018-5601-1. Epub 2018 Jul 6.

33. Patel BN, Rosenberg L, Willcox G, Baltaxe D, Lyons M, Irvin J, Rajpurkar P, Amrhein T, Gupta R, Halabi S, Langlotz C, Lo E, Mammarappallil J, Mariano AJ, Riley G, Seekins J, Shen L, Zucker E, Lungren M. Human-machine partnership with artificial intelligence for chest radiograph diagnosis. NPJ Digit Med. 2019 Nov 18;2:111. doi: 10.1038/s41746-019-0189-7. Erratum in: NPJ Digit Med. 2019 Dec 10;2:129. doi: 10.1038/s41746-019-0198-6

34. Verghese A, Shah NH, Harrington RA. What This Computer Needs Is a Physician: Humanism and Artificial Intelligence. JAMA. 2018 Jan 2;319(1):19–20. doi: 10.1001/jama.2017.19198

35. Jussupow E, Spohrer K, Heinzl A. Identity Threats as a Reason for Resistance to Artificial Intelligence: Survey Study With Medical Students and Professionals. JMIR Form Res. 2022 Mar 23;6(3):e28750. doi: 10.2196/28750

36. Castagno S, Khalifa M. Perceptions of Artificial Intelligence Among Healthcare Staff: A Qualitative Survey Study. Front Artif Intell. 2020 Oct 21;3:578983. doi: 10.3389/frai.2020.578983

37. Gille F, Jobin A, Ienca M. What we talk about when we talk about trust: theory of trust for AI in healthcare. Intell Based Med 2020. 1–2, 100001. doi:10.1016/j.ibmed.2020.100001

38. Liyanage H, Liaw ST, Jonnagaddala J, Schreiber R, Kuziemsky C, Terry AL, de Lusignan S. Artificial Intelligence in Primary Health Care: Perceptions, Issues, and Challenges. Yearb Med Inform. 2019 Aug;28(1):41–46. doi: 10.1055/s-0039-1677901. Epub 2019 Apr 25.

39. Huisman M, Ranschaert E, Parker W, Mastrodicasa D, Koci M, Pinto de Santos D, Coppola F, Morozov S, Zins M, Bohyn C, Koç U, Wu J, Veean S, Fleischmann D, Leiner T, Willemink MJ. An international survey on AI in radiology in 1,041 radiologists and radiology residents part 1: fear of replacement, knowledge, and attitude. Eur Radiol. 2021 Sep;31(9):7058–7066. doi: 10.1007/s00330-021-07781-5. Epub 2021 Mar 20.

40. Nori H, King N, McKinney SM, Carignan DM, Horvitz E. Capabilities of GPT-4 on Medical Challenge Problems. arXiv:2303.13375. 2023. 10.48550/arXiv.2303.13375

41. Thirunavukarasu AJ, Hassan R, Mahmood S, et al. (2023). How Can the Clinical Aptitude of AI Assistants Be Assayed? J Med Internet Res, 25, e51603. doi: 10.2196/51603

42. Magrabi F, Ammenwerth E, McNair JB, De Keizer NF, Hyppönen H, Nykänen P, Rigby M, Scott PJ, Vehko T, Wong ZS, Georgiou A. Artificial Intelligence in Clinical Decision Support: Challenges for Evaluating AI and Practical Implications. Yearb Med Inform. 2019 Aug;28(1):128–134. doi: 10.1055/s-0039-1677903. Epub 2019 Apr 25.

43. Daneshjou R, Smith MP, Sun MD, Rotemberg V, Zou J. Lack of Transparency and Potential Bias in Artificial Intelligence Data Sets and Algorithms: A Scoping Review. JAMA Dermatol. 2021 Nov 1;157(11):1362–1369. doi: 10.1001/jamadermatol.2021.3129.

44. Sendak MP, Gao M, Brajer N, Balu S. Presenting machine learning model information to clinical end users with model facts labels. NPJ Digit Med. 2020. 3, 41. doi:10.1038/s41746-020-0253-3

